# Factors affecting nurses’ duty to care during the COVID-19 pandemic

**DOI:** 10.1101/2021.10.21.21265272

**Authors:** Hyerine Shin, Kyung hee Kim, Ji-su Kim, Yeun-hee Kwak

## Abstract

**Background:** Although the demand for nursing care in disaster situations has grown, there has been a lack of discussion on nurses’ duty to care in these situations.

**Aim:** This study aimed to examine South Korean nurses’ duty to care during the COVID-19 crisis and identify factors influencing the same.

**Research design:** This was a cross-sectional descriptive research study, using a structured online questionnaire.

**Participants and research context:** Korean registered nurses (n = 342) in a clinical setting were recruited. Participants responded to a demographic questionnaire and the Nash Duty to Care Scale. After excluding missing values, data from 320 nurses were analyzed.

**Findings:** Older age and working at a general hospital increased nurses’ duty to care. Being male, higher education level, and working at a general hospital increased perceived risk. Older age, more clinical career experience, a master’s degree or above, and working at a higher-level hospital increased nurses’ confidence in their employer. Older age and higher monthly wage increased perceived obligation. Older age, job position, 3–7 years of clinical experience, working at the internal medicine department, and working at the tertiary hospital were factors associated with increased professional preparedness. Age was a factor influencing all dimensions of duty to care, except perceived risk. Clinical career influenced both confidence in employer and professional preparedness.

**Conclusion:** Given the lack of research on nurses’ duty to care, this study expands the scope of nursing research. In the future, more active research on nurses’ duty to care should be conducted.

## INTRODUCTION

The coronavirus disease-19 (COVID-19) pandemic has highlighted an ethical issue regarding healthcare workers. Centers for Disease Control and Prevention (CDC) reported that between February 12, 2020, and April 9, 2020, 19% of 315,531 confirmed COVID-19 cases were those of health care workers.[1] These results are not surprising as the three countries most severely affected by the Ebola pandemic reported that the viral infection rate among healthcare workers was 42 times that of the general public, and the mortality rate was twice as high.[2]

Frontline healthcare workers at the forefront of disaster situations take various risks in a highly strained environment to provide care for patients.[3] The concept of duty to care encompasses ethical aspects of providing care to patients even in situations where health care providers might be at risk themselves.[4] In the past, a “social contract model” was used to define duty to care describing it as a negotiation between the medical profession and society.[5] In modern times, as various types of disasters occur, the perspective on duty to care has also changed—nurses may experience ethical conflicts between human rights and patient wellbeing.[6] Healthcare workers, as professionals with trained skills, have a moral obligation to provide care for the continuation of the medical system, from responding to infectious diseases to providing medical services not related to infectious diseases.[7] However, in reality, since patients are both victims and carriers of disease, they are also dangerous to healthcare workers. Therefore, healthcare workers often face difficulties in the traditional patient-expert role.[8]

Especially, since Florence Nightingale, nurses have been an essential professional group in responding to disasters and mass accidents.[9,10] In disaster situations, nurses act as first responders,[11] playing an important role in improving the health of disaster victims and restoring resilience.[12] In general, disaster response systems are built with the assumption that an adequate number of personnel will be deployed at the disaster site. If fewer staffs participate in disaster response, the safety, quality, and sustainability of medical services provided will be at risk.[9] The shortage of medical personnel not only increases mortality due to missing early warning signs among patients, but also causes medical errors due to fatigue and increases the spread of infectious diseases.[4] These suggest the need for an investigation about the degree of nurses’ duty to care.

Previous research has focused on the construct of “duty to treat” that encompasses all medical, healthcare, and first aid workers, rather than the “duty to care” that only involves nurses. Moreover, previous studies on the duty of treatment confirmed the medical personnel’s’ “willingness to work” by focusing on the availability of the medical system, not the duty of treatment itself. Most research on nurses’ duty to care has been limited to confirming the concept through literature reviews or examining the perception of medical personnel who provide nursing and medical care in disaster situations through qualitative research. Regarding the duty to care of nurses, Kangasniemi et al.[6] pointed out that nurses’ rights include innate rights, such as human rights, rights under medical law, and professional ethics. Sokol[13] emphasized the need for a discussion on the limits of duty to care and argued that it cannot be enforced in a situation that exceeds limits drawn through social consensus. However, to our knowledge, no study has quantitatively analyzed this aspect.

Therefore, this study aims to bridge this gap by quantitatively analyzing the level of Korean clinical nurses’ duty to care during the COVID-19 crisis using Nash’s Duty to Care Scale (NDCS), and identifies the factors influencing the same.

## METHODS

This was a cross-sectional descriptive research study to examine Korean nurses’ duty to care and identify influencing factors. The study was conducted online, using a structured, self-report questionnaire. Data included nurses’ sociodemographic characteristics and responses to the NDCS. The collected data were analyzed using SPSS Statistics 25.0 program.

### Research participants and data collection

Participants were Korean registered nurses holding a license, currently working at a clinical nursing practice. Those who resigned, took a leave of absence, and did not work at clinical nursing were excluded. Using the G-power version 3.1.9.7 program, with significance level (α) .05, power (1-β) .95, and effect size (r) .25, the sample size was anticipated to be at least 280. Considering a dropout rate of 20%, 336 participants were targeted. Participants were recruited through a link posted in a banner advertisement of Nursescape (https://www.nurscape.net), which includes more than 330,000 Korean nurse subscribers, using the convenience sampling method. The purpose and method of the study were described in detail before the survey. Only those who voluntarily agreed to participate in the study completed the self-report questionnaire. A total of 342 nurses were recruited during the survey period. However, after excluding 22 questionnaires with missing data, analysis was conducted for data from 320 nurses.

### Data analysis

The collected data were analyzed using SPSS Statistics 25.0 program. The demographic characteristics of the participants were analyzed using descriptive statistics such as frequency, percent, mean, and standard deviation. The nurses’ duty to care was analyzed using descriptive statistics such as mean, standard deviation, maximum and minimum values. Nurses’ duty to care according to demographic characteristics were analyzed by independent t-test and one-way analysis of variance (ANOVA), followed by a post-test by Duncan. Stepwise multiple regression analysis was used to identify factors affecting nurses’ duty to care.

### Instruments

Nurses’ duty to care was measured using the Nash Duty to Care Scale (NDCS) developed by Nash.[14] NDCS includes 19 items across four subscales: Perceived risk (7 items, Cronbach’s α .91), Confidence in employer (3 items, Cronbach’s α .81), Perceived obligation (5 items, Cronbach’s α .83), and Profession preparedness (4 items, Cronbach’s α .85). Participants respond on a 5-point Likert scale (1 = strongly disagree; 5 = strongly agree). Negatively written items were calculated by converting them into inverse scores. Possible scores range from 19 to 95, with higher scores indicating more willingness to respond in a disaster situation. Two nursing professors, one English professor, and two nurses including researchers who are bilingual speakers fluent in Korean and English, translated and revised NDCS[14] into Korean. The developer ensured that the translated scale retained its original meaning. Several parts of the translated version were modified based on the developer’s direction. This study has used the final version of this scale after approval. In this study, the reliability of the tool was Cronbach’s α .78.

Participants responded to questions regarding sociodemographic characteristics referring to previous studies:[15-17] age, marital status, religion, gender, education level, job position, working department, monthly income, and the type of hospital (size of hospital, type of department, severity of patients treated). Based on Jang,[18] who revised and supplemented Benner’s[19] model according to the Korean population, careers were classified into 4 stages: the beginner level (up to 1 year of experience), advanced beginner level (more than 1 year and less than 3 years of experience), the competent level (more than 3 years and less than 7 years of experience), and the proficient level (7 years or more of experience).

### Ethical considerations

This study was approved by the Institutional Review Board (IRB) of [blinded for review] University (approval no.: 1041078-202103-HRBM-080-01). Informed consent was obtained from all participants prior to the study. Participants were guaranteed anonymity, were informed that all information provided by them would only be used for research purposes, and that participation in the study was voluntary. All information collected for research would be safely disposed of in a non-recoverable manner after the retention period is over.

## RESULTS

Participants (n = 320) were registered nurses from Korea with an average age was 31.87 years. Most participants were female, unmarried, bachelor’s degree holders, staff nurses, and were not religious (Table 1). Nurses’ average score of duty to care was 62.15. Average scores on subscales were 24.13, 8.62, 18.12, and 11.29, for Perceived risk, Confidence in employer, Perceived obligation, and Profession preparedness, respectively (Table 2).

**Table 1.** Sociodemographic characteristics of nurses

| Characteristics | Categories | N (%) or M±SD |
| --- | --- | --- |
| Age |  | 31.87±8.25 |
| Marital status | Single | 241 (75.3) |
|  | Married | 76 (23.8) |
|  | Other | 3 (0.9) |
| Religion | Yes | 119 (37.2) |
|  | No | 201 (62.8) |
| Sex | Female | 307 (95.9) |
|  | Male | 13 (4.1) |
| Education | Associate degree | 35 (10.9) |
|  | Baccalaureate | 251 (78.4) |
|  | Master or above | 34 (10.6) |
| Length of service (years) | < 1 | 68 (21.3) |
|  | ≥1 to < 3 | 65 (20.3) |
|  | ≥3 to < 7 | 108 (33.8) |
|  | ≥7 | 79 (24.7) |
| Job position | Staff nurse | 286 (89.4) |
|  | Charge nurse | 21 (6.6) |
|  | Head nurse or above | 13 (4.1) |
| Work unit | Internal Medicine | 80 (25.0) |
|  | General Surgery | 87 (27.2) |
|  | Special Unit | 72 (22.5) |
|  | Others | 81 (25.3) |
| Monthly Wage<br>(10,000 Won) | < 300 | 141 (44.1) |
|  | ≥300 | 179 (55.9) |
| Type of Hospital | Tertiary Hospital | 151 (47.2) |
|  | General Hospital | 101 (31.6) |
|  | Other hospitals | 68 (21.3) |

**Table 2.** Nurses’ duty to care in disaster situations

| Characteristics | M±SD | Min | Max |
| --- | --- | --- | --- |
| Total Score | 62.15±8.71 | 30.00 | 89.00 |
| Perceived Risk | 24.13±4.19 | 12.00 | 35.00 |
| Confidence in Employer | 8.62±2.73 | 3.00 | 15.00 |
| Perceived Obligation | 18.12±3.28 | 8.00 | 25.00 |
| Professional Preparedness | 11.29±3.58 | 4.00 | 20.00 |

**Table 3.**
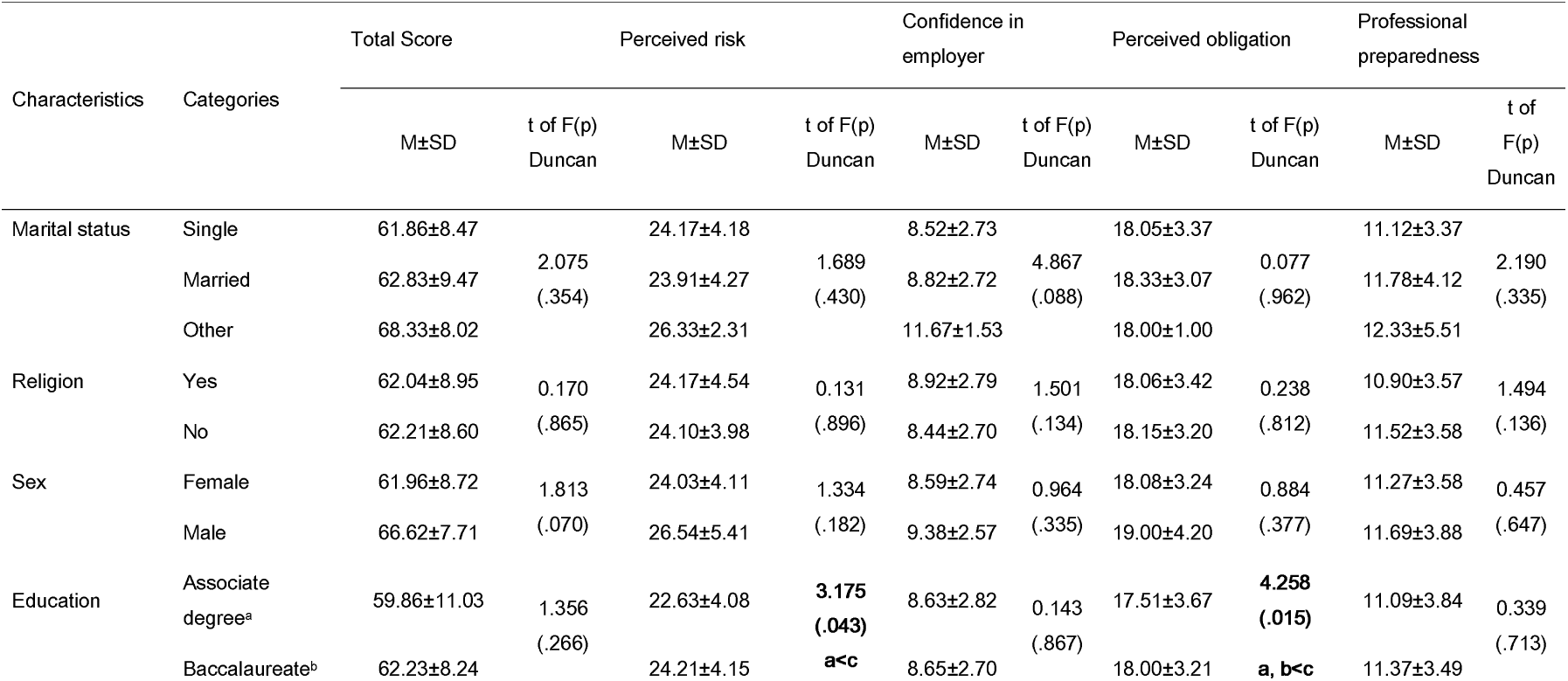
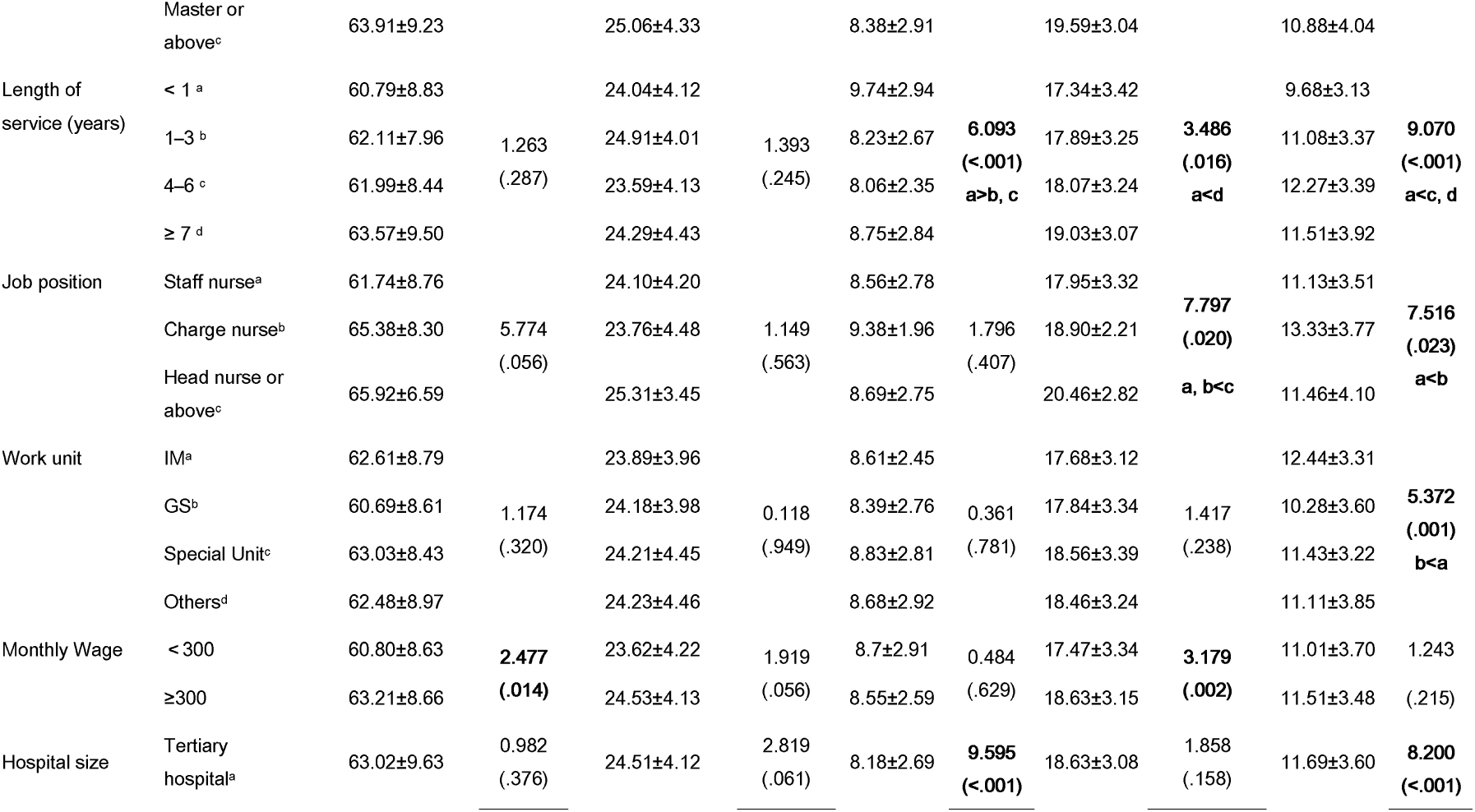
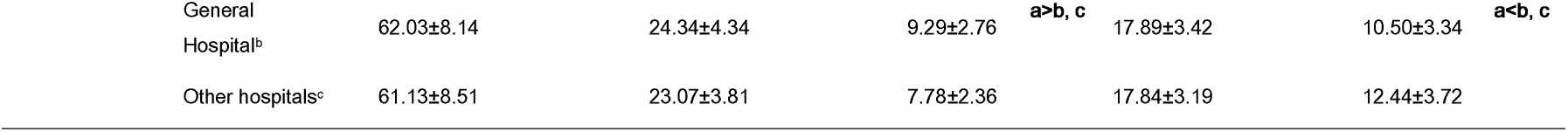
Differences in duty to care during disaster situations based on sociodemographic characteristics

**Table 4.**
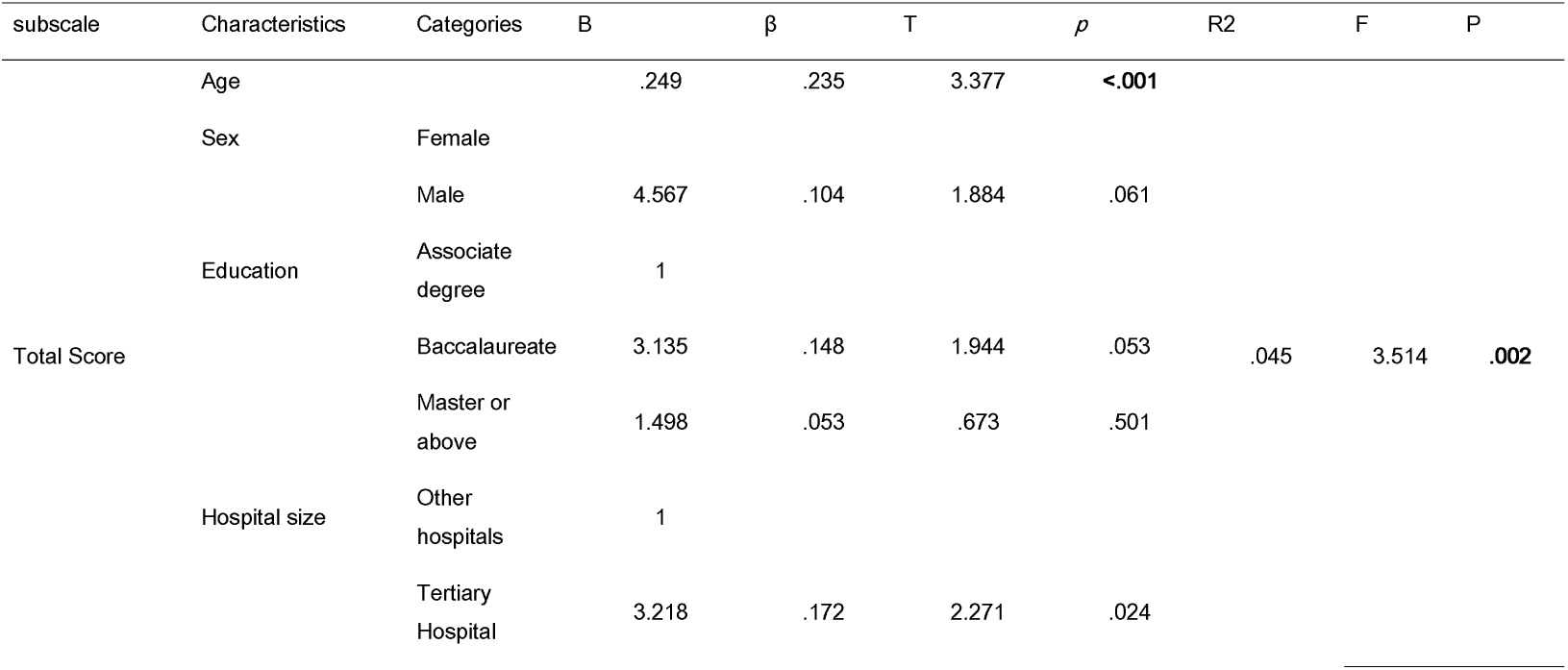
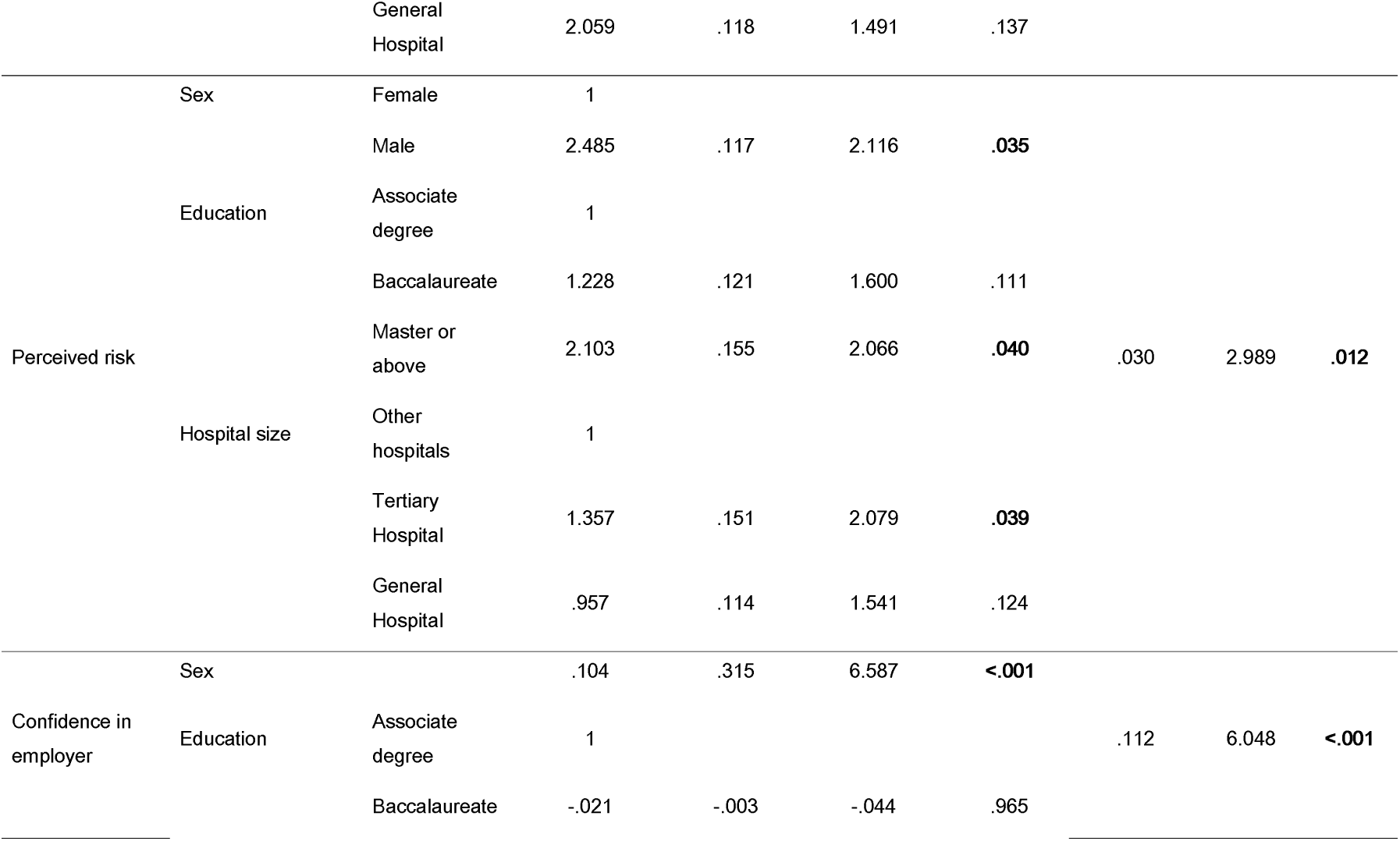
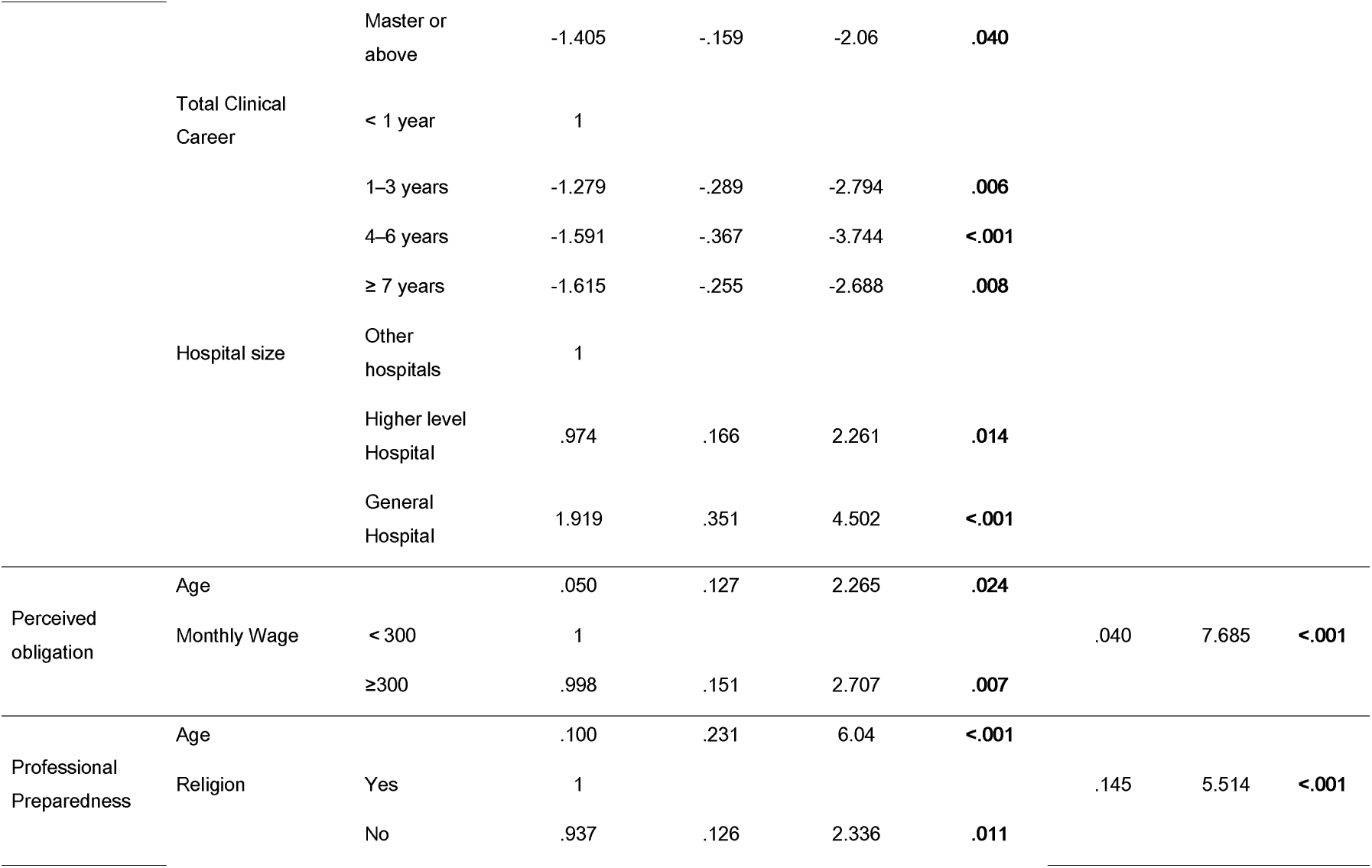
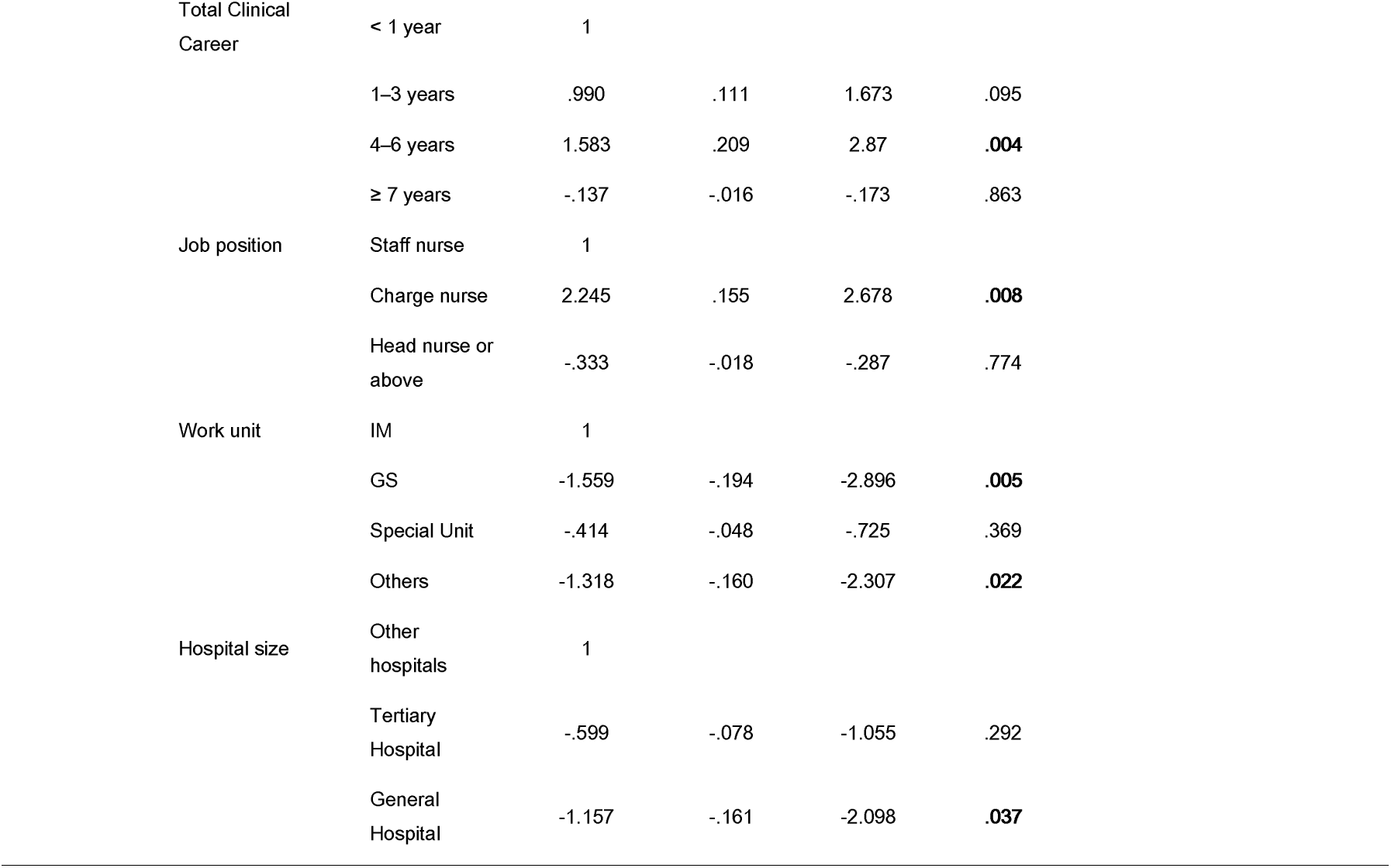
Multiple regression analysis of factors affecting duty to care

The results of t-test and ANOVA indicate that perceived risk was higher among nurses with a master’s degree or higher (F = 3.175) compared with those who held a bachelor’s degree (F = 0.043). Confidence in employer was significantly higher among nurses with less than 1 year of experience than those with 1–7 years of experience (F = 6.093, p < 0.001). When working at a tertiary general hospital, nurses’ confidence in employer was significantly higher than when working at a general hospital and other hospitals (F = 9.595, p < 0.001).

Perceived obligation showed a significant difference according to education level, total clinical career, position, and monthly wage. A master’s degree or higher yielded significantly higher scores in the confidence in employer dimension, compared with an associate or bachelor’s degree (F = 4.258, p = 0.015). Participants with 7 or more years of experience had significantly higher scores than those with less than 1 year of experience (F = 3.486, p = 0.016). In addition, nurses with job position as head nurses or above (F = 7.797, p = 0.02) had higher scores than staff nurses (F = 7.797, p = 0.02).

Professional preparedness indicated significant differences based on total clinical career, position, work unit, and type of hospital. Nurses with less than 1 year of experience reported lower professional preparedness than that of those with 3–7 years and 7 or more years of experience (F = 9.07, p < 0.001). Internal medicine unit reported higher professional preparedness than the general surgery unit (F = 5.372, p = 0.001). Moreover, working at tertiary hospitals indicated lower professional preparedness than working in general hospitals and other hospitals (F = 8.200, p < 0.001). Duty to care was significantly higher among those who received 3 million won or more as monthly wages, than those who received less than 3 million won (t = 2.477, p = 0.014).

To verify the factors affecting duty to care, a stepwise multiple regression analysis was performed by inputting sociodemographic characteristics as independent variables. Factors influencing perceived risk were being male (β = .117, p = .035), holding a master’s degree or higher (β = .155, p = .04), and working at tertiary hospital (β = .151, p = .039). Factors affecting confidence in employer were older age (β = .315, p < .001), holding a master’s degree or higher (β = .159, p = .040), 1–3 years of work experience (β = .289, p = .006), 3–7 years of experience (β = .367, p < .001), 7 or more years of experience (β = .255, p = .008), working at a tertiary hospital (β = .166, p = .014) and general hospital (β = .351, p < .001). Factors affecting perceived obligation were older age (β = .127, p = .024) and monthly wage of 3 million won or more (β = .151, p = .007). Factors influencing professional preparedness were older age (β = .231, p < .001), being not religious (β = .126, p = .011), 3–7 years of work experience (β = .209, p = .004), working as a charge nurse (β = .155, p = .008), being a part of the general surgery unit (β = .194, p = 0.005) or other unit (β = .160, p = .022), and working at a tertiary hospital (β = .161, p = .037). Overall, the factor influencing duty to care the most was older age (β = .235, p < .001).

Over duty to care (F = 3.514, p = 0.002) and its sub-scales had suitable regression models. It was confirmed that there was no autocorrelation between each independent variable. As a result of examining the multicollinearity between the independent variables, the Variances Inflation Factor (VIF) values for all variables ranged from the minimum value of 1.005 and the maximum value of 1.738 to less than 10, indicating that there was no problem with multicollinearity between the variables.

## DISCUSSION

This study examines Korean nurses’ duty to care during the COVID-19 pandemic and identifies factors influencing the same. Participants in this study reported an average duty to care score of 62.15 (Table 2), which is lower than previous study[20] that measured duty to care among Taiwanese and US nurses. Particularly, scores on subscales of confidence in employer, perceived obligation, and professional preparedness yielded lower scores compared with nurses in Taiwan and the United States (Table 2). This may be because nurses in this study were working, directly or indirectly, during the prolonged COVID-19 pandemic. At the beginning of the pandemic, the dedication of nurses in Korea was lauded as a “heroic act.” However, adequate support was not provided, leading to most nurses working without the education and training of disaster nursing.[21] Therefore, dissatisfaction with insufficient support and resources may have lowered Korean nurses’ duty to care.

Duty to care increased with older age. This is consistent with previous studies, showing that nurses aged 35 or older had increased will and ability to go to work in disaster situations.[22] Gender, education, and hospital size were factors influencing perceived risk. This accords with previous studies where males were more likely to take risks for patients than females.[23,24] In most cultures, there are the traditional expectations for women to nurture children more than men,[24] and that women are more likely to avoid risks that may threaten them and their children. Perceived risk among nurses working in tertiary hospitals was higher than that of other hospitals. Although there are some limits to comparison due to systemic differences, it may be due to burnout. Usually, small- and medium-sized hospitals in Korea have much lower wages, poor welfare, chronic shortage of nurses, and more additional tasks such as guidance and supervision of non-professionals, than tertiary hospitals.[25] Considering that burnout lowers the level of nursing professionalism and vocational awareness,[26] burnout caused by the relatively poor working environment of other hospitals may reduce the nurses’ will to care in disaster situations.

Confidence in employer increased with older age, while it showed a tendency to gradually decrease as the level of education and experience increased. As careers and educational backgrounds flourish, the expectations about the working environment and organization grow. As support from an organization is associated with trust,[27-29] dissatisfaction from the working environment and organization may lower the confidence in employer. In particular, 3 to 7 years of experience was found to be the greatest factor influencing confidence in employer. The nurses with 3 to 7 years of experience are at the competent stage, proficient with nursing practice, grow as professionals, and play the role of a preceptor for novice nurses.[18] Therefore, to improve duty to care, it is necessary to increase confidence in employer for nurses who are in the competent stage (3 to 7 years). This also suggests that it is necessary to develop a customized organizational trust promotion program for each career.

Perceived obligation increased as the age and monthly wage increased. This result is consistent with previous studies, indicating that the higher the income, the higher the sense of calling.[30,31] Similar to the concept of perceived obligation, sense of calling refers to the recognition of accepting work itself as the purpose and meaning of an individual’s life.[32,33] According to Dobrow, sense of calling is not innate and can be changed over time, due to various factors.[34] The monetary reward such as monthly salary is a result of individual performance and at the same time, serves as a basis for sense of calling, and may increase perceived obligation.

Professional preparedness tended to increase with older age and was higher among those with 3 to 7 years of experience than those with less than one year of experience. These results are partially consistent with previous studies which state that, as age and experience increase, various clinical careers can be built, which can have a positive effect on disaster nursing competency.[35] Moreover, compared with the internal medicine unit, professional preparedness was lower in the general surgery unit and other units. In Korea, epidemics of infectious diseases such as Middle East Respiratory Syndrome (MERS), Severe Acute Respiratory Syndrome (SARS), and COVID-19 have occurred more frequently than national mass traumatic disasters, such that internal medicine units have participated in disaster situations more than others. The experience of participating in disaster nursing increases its capacity.[36] This may have increased the IM nurses’ professional preparedness. Position appeared to be the greatest influencing factor for professional preparedness, which is different from previous studies.[37-39] Charge nurses are practitioners in the intermediate stage between the head and the staff nurse and are involved in the work of both nursing practice and nursing management.[40] In disaster nursing, the roles of nursing managers who direct and supervise nursing practices for disaster victims and disaster practice nursing sites may differ, and accordingly, the required disaster nursing competency may also appear differently. Therefore, it is necessary to study disaster nursing competency considering the role characteristics of the charge nurse in the future.

Overall, sociodemographic characteristics explained confidence in employer (Adjusted R2 = .112) and professional preparedness (Adjusted R2 = .145) relatively well, among the sub-scales of duty to care. These results suggest that an intervention considering the sociodemographic characteristics is needed to enhance nurses’ duty to care, especially regarding confidence in employer and professional preparedness.

This study has some limitations. First, since this study was conducted through an online survey, it is necessary to be careful about generalizing the findings as there may be a selection bias of users who use the Internet. Second, due to the lack of prior research, only sociodemographic characteristics were analyzed to explain nurses’ duty to care. However, this study is the first to quantitatively analyze Korean nurses’ duty to care, using the translated version of NDCS. Therefore, findings from this study may be used as basic data for related studies in the future. Moreover, this study included nurses practicing in the clinic during the COVID-19 pandemic. This is meaningful in that it served as an opportunity to understand duty to care of nurses close to the clinical field.

## CONCLUSION

This study is the first cross-sectional study in Korea to measure nurses’ duty to care through a structured survey and to analyze factors influencing the same. Nurses have an important role in responding to disaster victims at the frontline. However, nurses are facing a large-scale pandemic without considering their duty to care. This has led to various ethical issues. Nurses’ willingness and effort to provide care in disaster situations is an important component of disaster response. Therefore, it is crucial to improve this through various efforts and interventions. These efforts must be made socially and policy-wise, beyond the individual or organizational level. In the future, various efforts are needed to expand and analyze related factors through active research nurses’ duty to care and to reinforce them.

## Supporting information

Supplemental title page

cover letter

## Data Availability

All data produced in the present study are available upon reasonable request to the authors

