## Supplementary material for "Factors affecting nurses’ duty to care during the COVID-19 pandemic": cover letter

[October 19, 2021]

John McMillan

Editor-in-Chief

*Journal of Medical Ethics*

Dear Editor:

I wish to submit an empirical paper for publication in the *Journal of Medical Ethics*, titled “Factors affecting nurses’ duty to care during the COVID-19 pandemic.” The paper was coauthored by Hyerine Shin, Ji-su Kim, and Yeun-hee Kwak.

This study examines Korean nurses’ duty to care during the COVID-19 pandemic and identifies factors influencing the same. We believe that our study makes a significant contribution to the literature because it is the first to quantitatively analyze Korean nurses’ duty to care, using the translated version of Nash Duty to Care Scale.

Further, we believe that this paper will be of interest to the readership of your journal because our findings recognize the need of an intervention considering the socio-demographic characteristics to enhance nurses’ duty to care, especially regarding confidence in employer and professional preparedness.

This manuscript has not been published or presented elsewhere in part or in entirety and is not under consideration by another journal. All study participants provided informed consent, and the study design was approved by the appropriate ethics review board. We have read and understood your journal’s policies, and we believe that neither the manuscript nor the study violates any of these. There are no conflicts of interest to declare.

Thank you for your consideration. I look forward to hearing from you.

Sincerely,

Kyung hee Kim

Department of Nursing, Chung-Ang University, 84 Heukseok-ro Dongjak-Gu, 06974,

Seoul, Republic of Korea
